# Trends in occupational respiratory conditions with short latency from 1999 to 2019 in the UK - evidence from the Surveillance of Work-related and Occupational Respiratory Disease (SWORD) reporting scheme

**DOI:** 10.1101/2023.05.19.23290195

**Authors:** Ana Barradas, Ireny Iskandar, Melanie Carder, Matthew Gittins, Laura Byrne, Susan Taylor, Sarah Daniels, Ruth E. Wiggans, David Fishwick, Martin Seed, Martie van Tongeren

**Affiliations:** Centre for Occupational and Environmental Health, Faculty of Biology, Medicine and Health, University of Manchester, 4^th^ Floor, Block C, Ellen Wilkinson Building, Oxford Road, Manchester M13 9PL; Department of Biostatistics, Division of Population Health, Health Services Research & Primary Care, University of Manchester, Oxford Road, Manchester M13 9PL; Chest Clinic, Wythenshawe Hospital, Manchester, UK; Centre for Workplace Health, Sheffield Teaching Hospitals NHS Foundation Trust, Sheffield, UK

**Keywords:** Occupational health, lung/respiratory disease, short latency, surveillance scheme, SWORD

## Abstract

**Background:** Occupational short-latency respiratory disease (SLRD; predominantly asthma, rhinitis, hypersensitivity pneumonitis, and occupational infections) prevalence is difficult to determine but certain occupations may be associated with increased susceptibility.

**Aims:** This study aimed to examine which occupations and industries are currently at high risk for SLRD and determine their respective suspected causal agents based on cases reported by physicians to the Surveillance of Work-related and Occupational Respiratory Disease (SWORD) scheme in the UK.

**Methods:** SLRD cases reported to the SWORD scheme between 1999 and 2019 were analysed to determine directly standardised rate ratios (SRR) by occupation against the average rate for all other occupations combined.

**Results:** Bakers and flour confectioners showed significantly raised SRR for occupational rhinitis (234.4 [95% CI, 200.5 - 274.0]) and asthma (59.9 [95% CI, 51.6 - 69.5]). Chemical and related process operatives also presented raised SRR values for these two conditions, with SRR of 29.5 [95% CI, 24.3 - 35.7] and 21.0 [95% CI, 16.9 - 26.1] for rhinitis and asthma, respectively. SRR were also significantly raised for vehicle spray painters when considering occupational asthma (63.5 [95% CI, 51.5 - 78.3]) alone, and laboratory technicians were also amongst the top three increased SRR for rhinitis (18.7 [95% CI, 15.1 - 23.1]). The suspected agents most frequently associated with these occupations and conditions were flour, isocyanates, and laboratory animals and insects. Metal machining setters and setter-operators showed increased SRR for occupational hypersensitivity pneumonitis (42.0 [95% CI, 29.3 - 60.3]), largely due to cutting/soluble oils. The occupation mostly affected by infectious disease was welding trades (12.9 [95% CI, 5.7 - 29.3]) and the suspected causal agent predominantly reported for this condition was pathogens and microorganisms, with a predominance of *Mycobacterium tuberculosis*.

**Conclusions:** This study identified the occupational groups at increased risk of developing a SLRD based on data recorded over a recent two-decade period in the UK. Asthma and rhinitis were identified as the prevailing conditions and hypersensitivity pneumonitis as a potentially rising respiratory problem in the metalworking industry.

## Introduction

Respiratory illnesses caused by harmful inhaled occupational exposures are diverse in nature (1) and exhibit varying latent periods; the period between first exposure to the causative agent and the development of the condition. For example, dust related diseases such as coal workers’ pneumoconiosis or silicosis, and occupational cancers, are generally associated with longer latent periods (2, 3). However, many occupational respiratory illnesses are associated with shorter latent periods, and include occupational rhinitis and asthma, hypersensitivity pneumonitis and certain infections.

These short latency respiratory diseases (SLRDs) are considered to represent a group of illnesses, caused by workplace exposures, where the host response to the causative agent develops more rapidly and are associated with a wide variety of exposure types (4). A significant proportion of these conditions are either allergic or immunologically mediated (5, 6) on the background of high levels of allergic disease in general populations (7) or infectious in origin. Whilst having less effects on mortality, SLRDs are evidently associated with significant morbidity, job loss (8) and wider socioeconomic impacts (9).

The UK based SWORD scheme (Surveillance of Work-related and Occupational Respiratory Disease) is a national reporting system for physicians that permits collation of cases of occupational lung disease (10), along with contextual information that includes co-existing diagnoses, likely causative exposure, occupation and work sector.

Given that comparatively little is reported about shorter latency conditions from reporting schemes (11-14), with perhaps the exception of occupational asthma, and the comparative lack of data related to both occupational rhinitis (15, 16) and infections (17), we chose to report here information related to all such cases reported to SWORD over the period 1999 to 2019.

## Methods

Cases of occupational SLRD reported to SWORD (18, 19) between 1999 and 2019, were used for the purposes of this analysis. Cases were reported anonymously by respiratory physicians in the UK; either as a core or sample reporter. Core reporters return all cases of occupational respiratory disease diagnosed monthly throughout the year, while sample reporters return only cases diagnosed during a single randomly selected month per year.

Information included in the report to SWORD includes age, gender, occupation, industry, first half of postcode, and up to three suspected causal agents attributed to the case. Data are coded according to their respective system classifications; occupation by the Standard Occupational Classification (SOC) 2010 (20), industry of work by the Standard Industrial Classification (SIC) (21), and an in-house causal agent classification system is used to collate exposure type. A total estimated number of cases per year was calculated by adding the actual reported cases from the core reporters to the number of cases reported by the sample reporters factored by 12. The methodologies adopted have previously been described in more detail (18).

Cases extracted for this work included (i) asthma, (ii) rhinitis, (iii) hypersensitivity pneumonitis and (iv) infectious diseases. All other cases were also assessed for completeness, and a priori categorised as (v) “other”. All short latency disease cases for which there was a co-diagnosis with a long-latency disease were discarded, as well as those for which the causal agent was specified as asbestos. Inhalation accidents (22), bronchitis/emphysema (COPD), non-malignant pleural disease (predominantly plaques, predominantly diffuse, and asbestos related pleural effusion), mesothelioma, lung cancer, pneumoconiosis (asbestosis, silicosis, and coal workers’ pneumoconiosis), and unspecified cases, which were unclear about the latency were all excluded from this analysis. Data from 2019 onward were excluded from these calculations due to the atypical reporting pattern during the COVID-19 pandemic.

Directly standardised rate ratios (SRR) per occupation were calculated for all relevant diagnostic categories. For each occupation, the incidence rate of a particular diagnosis (or individual diagnostic category) was obtained by averaging the number of total cases of the individual occupations weighted by their relative sizes according to employment-related statistics in the UK (23). Considering that the population distribution is not homogeneous regarding occupational groups, and to allow for a meaningful comparison between groups of different professional occupations, rates for each occupation were directly standardised by the average rate of all other occupations combined. Approximate 95% confidence intervals (CI) were estimated using a Taylor series variance estimator to account for the fluctuation inherent to the weighting of sample reports (24), and a finite population correction factor of 0.3 was used to adjust for the proportion of eligible chest physicians that report to SWORD (25).

The Health and Occupation Research (THOR) network (including EPIDERM and SWORD) has National Health Service ethics approval given by the Northwest (Haydock) Research Ethics Committee (22/NW/0082).

## Results

A total of 3476 cases (core plus unweighted sample cases) were reported over the study time period. Table 1 shows the number of actual and estimated cases and diagnoses by each disease category. Overall, cases were predominantly reported in men (71%) with a mean age of 43 ± 12.2 (SD) years, and predominantly reported in the manufacturing industry (54.3%), particularly in the manufacture of motor vehicles, trailers and semi-trailers (22.0%) and the manufacture of food products (19.3%). Next most frequent industrial sectors were professional, scientific and technical activities (8.7%), where the majority of cases (86.4%) were within scientific research and development.

**Table 1.** Number and percentage of short latency respiratory disease (SLRD) cases reported by chest physicians to SWORD^*^ (1999-2019). Mean age and sex are presented for all actual cases (core plus unweighted sample cases).

| Disease category | Actual cases<sup>1</sup><br>(% column) | Estimated cases<sup>2</sup><br>(% column) | Age<sup>3</sup> (mean ± SD)<br>(years) | Sex<sup>3</sup> |  |
| --- | --- | --- | --- | --- | --- |
|  |  |  |  | Female (% row) | Male (% row) |
| **Asthma** | **2784** (75%) | **5941** (73%) | **44 ± 11.7** | **770** (28%) | **2011** (72%) |
| **Rhinitis** | **588** (16%) | **731** (9%) | **36 ± 10.6** | **189** (32%) | **399** (68%) |
| **Hypersensitivity pneumonitis** | **209** (6%) | **770** (10%) | **52 ± 12.1** | **46** (22%) | **163** (78%) |
| **Infectious disease** | **72** (2%) | **501** (6%) | **43 ± 12.8** | **30** (42%) | **41** (57%) |
| **Other SLRD** | **80** (2%) | **157** (2%) | **48 ± 11.5** | **26** (33%) | **54** (68%) |
| **Total diagnoses<sup>4</sup>** | **3733** | **8100** | **43 ± 12.2** | **1061** (28%) | **2668** (71%) |
| **Total cases** | **3476** | **7818** | **44 ± 12.1** | **987** (28%) | **2485** (71%) |

<sup>1</sup>Actual cases refer to cases reported by core reporters plus unweighted cases reported by sample reporters
<sup>2</sup>Estimated cases refer to cases reported by core reporters plus 12 x cases reported by sample reporters
<sup>3</sup>Refers to cases where patient age/sex were reported
<sup>4</sup>Diagnoses refer to all disease categories reported per case
SD – standard deviation

The most reported occupations were skilled trades occupations (34.2%) and process, plant and machine operatives (25.3%). Of the former, the commonest occupational groups were food preparation trades (27.0%) and metal machining, fitting and instrument making trades (20.7%). Of the latter category, 47.3% were process operatives and 28.1% assemblers and routine operatives.

There were major differences in the incidence of SLRD between occupations, with the highest incidences seen in bakers and flour confectioners (SRR 58.9 [95% CI, 51.8 - 66.9]) and vehicle spray painters (SRR 48.9 [95% CI, 39.7 - 60.2]). Although still significantly raised, all other occupations had considerably lower incidence rates than these two occupations, in general. For example, metal working production and maintenance fitters (SRR 2.2 [95% CI, 1.8 - 2.8]), nurses (SRR 1.9 [95% CI, 1.5 - 2.3]), and cleaners and domestics (SRR 1.5 [95% CI, 1.2 - 1.9]) showed the lowest SLRD incidence rates (Figure 1).

**Figure 1.**
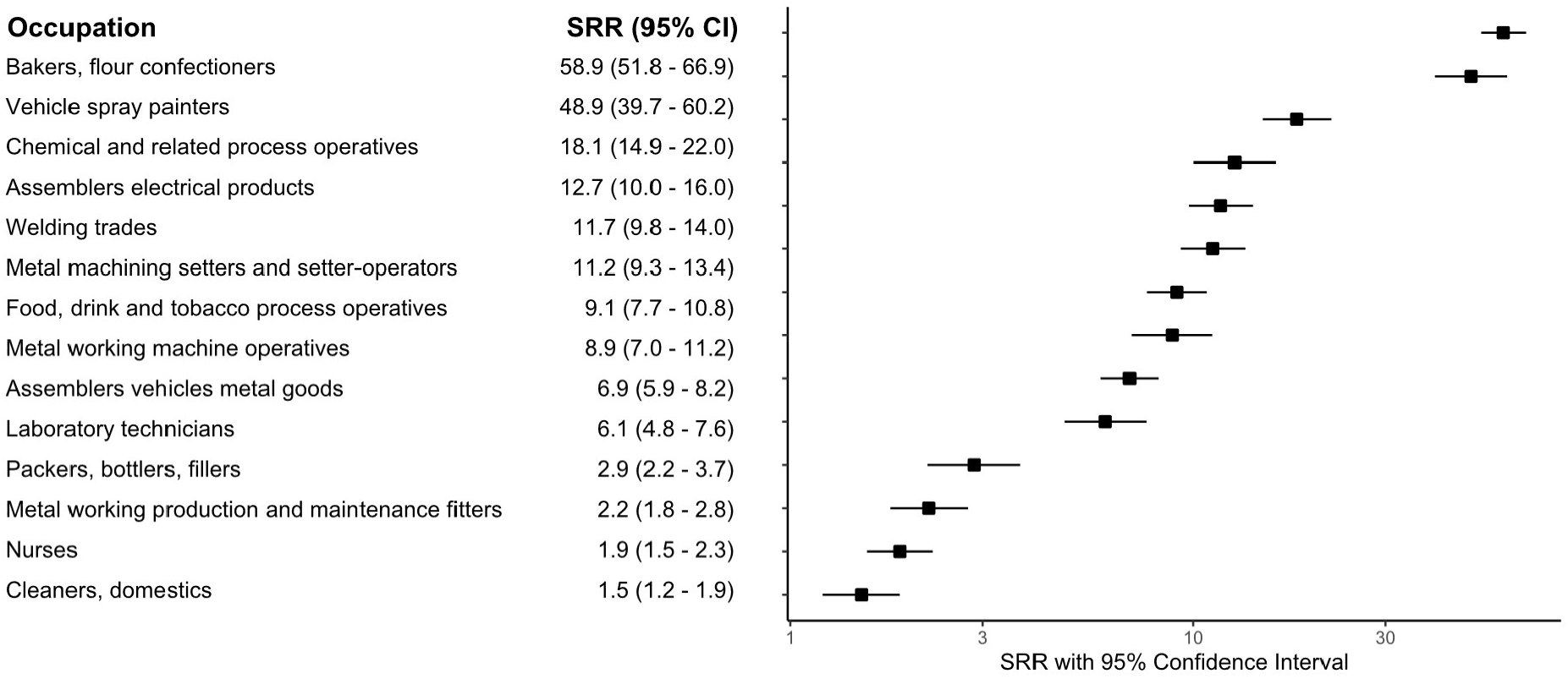
Standardised rate ratios (SRR) and 95% CI of medically reported occupational SLRD incidence reported to SWORD compared with all other employment sectors combined (1999-2019). SWORD, Surveillance of Work-Related and Occupational Respiratory Disease.

Asthma was the most frequently reported condition (2784 actual cases), comprising about 75% of all actual cases, followed by rhinitis (16%). Rhinitis was also the most commonly reported co-diagnosis, usually alongside asthma. Asthma cases were mostly male (72%), with a mean age of 44 ± 11.7 (SD) years, and with a predominance in the manufacture of food products and manufacture of motor vehicles, trailers and semi-trailers industries. Occupations with the highest incidence rates were bakers and flour confectioners (SRR 59.9 [95% CI, 51.6 - 69.5]), vehicle spray painters (SRR 63.5 [95% CI, 51.5 - 78.3]), and chemical and related process operatives (SRR 21.0 [95% CI, 16.9 - 26.1]) (Table 2). Isocyanates (24.2%) and flour (18.8%) were the most important agents causing occupational asthma, shown amongst other data relating to all other diagnoses (Figure 2).

**Table 2.** Standardised rate ratios (SRR) and 95% CI of medically reported occupational SLRD incidence reported to SWORD^*^ compared with all other employment sectors combined per diagnostic category (1999-2019).

| Occupation | SRR (95% CI) |  |  |  |
| --- | --- | --- | --- | --- |
|  | Asthma | Rhinitis | Hypersensitivity pneumonitis | Infectious disease |
| Bakers, flour confectioners | **59.9** (51.6 – 69.5) | **234.4** (200.5 – 274.0) | **13.1** (4.4 – 38.6) | - |
| Vehicle spray painters | **63.5** (51.5 – 78.3) | - | **1.3** (0.4 – 3.8) | - |
| Chemical and related process operatives | **21.0** (16.9 – 26.1) | **29.5** (24.3 – 35.7) | **0.6** (0.2 – 1.8) | - |
| Welding trades | **13.0** (10.8 – 15.7) | **1.0** (0.5 – 2.2) | **6.8** (2.4 – 18.8) | **12.9** (5.7 – 29.3) |
| Assemblers electrical products | **16.4** (12.9 – 20.8) | **1.9** (0.8 – 4.4) | - | - |
| Metal machining setters and setter-operators | **9.5** (7.6 – 11.7) | **3.7** (2.4 – 5.6) | **42.0** (29.3 – 60.3) | - |
| Food, drink and tobacco process operatives | **10.0** (8.3 – 12.0) | **14.1** (9.3 – 21.4) | **1.1** (0.6 – 1.9) | **8.6** (3.9 – 18.7) |
| Metal working machine operatives | **9.0** (6.9 – 11.6) | **2.7** (1.7 – 4.5) | **17.8** (10.1 – 31.3) | - |
| Assemblers vehicles metal goods | **8.0** (6.7 – 9.6) | **5.6** (3.8 – 8.2) | **9.6** (6.9 – 13.3) | - |
| Laboratory technicians | **5.6** (4.2 – 7.4) | **18.7** (15.1 – 23.1) | - | **8.7** (2.9 – 25.8) |
| Packers, bottlers, fillers | **2.1** (1.6 – 2.8) | **2.6** (1.8 – 3.7) | **4.2** (1.9 – 9.2) | **10.2** (4.7 – 22.2) |
| Metal working production and maintenance fitters | **2.3** (1.8 – 2.8) | **0.7** (0.4 – 1.2) | **3.9** (1.8 – 8.3) | **0.2** (0.1 – 0.5) |
| Nurses | **1.3** (1.1 – 1.6) | **1.6** (0.8 – 3.1) | **0.9** (0.3 – 2.5) | **10.0** (6.4 – 15.7) |
| Cleaners, domestics | **1.8** (1.4 – 2.2) | **0.1** (0.0 – 0.2) | **0.4** (0.3 – 0.7) | **1.3** (0.4 – 3.7) |

\*SWORD, Surveillance of Work-Related and Occupational Respiratory Disease

**Figure 2.**
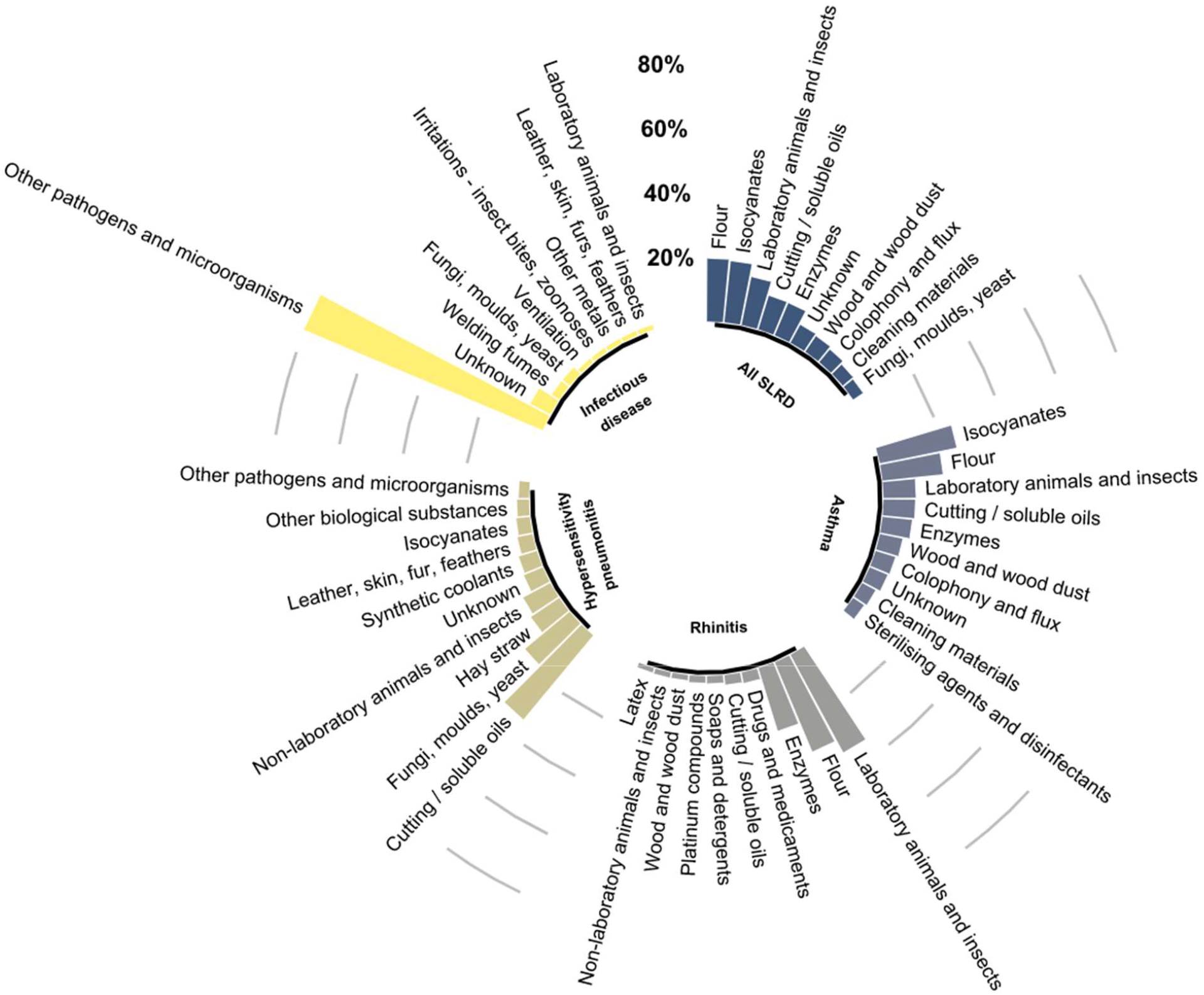
Suspected causal agents associated with the different occupational short latency respiratory disease (SLRD) categories reported to SWORD (1999-2019). The suspected agents subcategories are not necessarily grouped or ranked according to the hierarchy of the coding system. SWORD, Surveillance of Work-Related and Occupational Respiratory Disease.

Rhinitis cases (588 actual cases) were also mostly male (68%), with a mean age of 36 ± 10.6 (SD) years. Bakers and flour confectioners (SRR 234.4 [95% CI, 200.5 - 274.0]) showed the highest incidence of rhinitis, indicating that there is a significantly higher risk of developing rhinitis than of developing asthma. There was also a suggestion that chemical and related process operatives (SRR 29.5 [95% CI, 24.3 - 35.7]) and laboratory technicians (SRR 18.7 [95% CI, 15.1 - 23.1]) are occupations with an increased risk of occupational rhinitis. The incidence of rhinitis was significantly lower in metal working production and maintenance fitters (SRR 0.7 [95% CI, 0.4 - 1.2]) and cleaners and domestics (SRR 0.1 [95% CI, 0.0 - 0.2]) than in all other occupations combined (Table 2). Laboratory animals and insects (34.4%) and flour (30.1%) were the most important causes of rhinitis (Figure 2).

Hypersensitivity pneumonitis cases (209 actual cases) were again predominantly male (78%), with a mean age of 52 ± 12.1 years, as shown in Table 1. Metalworking industry activities such as metal machining setters and setter-operators (SRR 42.0 [95% CI, 29.3 - 60.3]) and metal working machine operatives (SRR 17.8 [95% CI, 10.1 - 31.3]) were the most affected occupations. These were followed by grain-based food industries, represented by bakers and flour confectioners (SRR 13.1 [95% CI, 4.4 - 38.6]). Occupations such as chemical and related process operatives (SRR 0.6 [95% CI, 0.2 - 1.8]), nurses (SRR 0.9 [95% CI, 0.3 - 2.5]), and cleaners and domestics (SRR 0.4 [95% CI, 0.3 - 0.7]) showed a significantly lower incidence of hypersensitivity pneumonitis than all other occupations, again as shown in Table 2. Metalworking fluids, such as cutting and soluble oils, were the main attributed causal agent in hypersensitivity pneumonitis (33.9% of all cases), followed by exposure to fungi, moulds and yeast (18.0%) (Figure 2).

Infectious disease was the diagnostic category with the lowest number of reported cases (72 actual cases), and the highest female proportion (42%). Welding trades (SRR 12.9 [95% CI, 5.7 - 29.3]), packers, bottlers, fillers (SRR 10.2 [95% CI, 4.7-22.2]) and nurses (SRR 10.0 [95% CI, 6.4 - 15.7]) ranked as the top three occupations at the highest risk of being reported as a case and only metal working production and maintenance fitters (SRR 0.2 [95% CI, 0.1 - 0.5]) showed a significantly lower incidence of infectious disease than all other occupations (Table 2). As expected, pathogens and microorganisms were the predominant suspected cause of infectious diseases (80.3%) (Figure 2). In particular, *Mycobacterium tuberculosis* (53%), other species of bacteria (43%) and fungi (2%) were attributed as causes of individual cases. More information is included in supplementary Figure S1.

Figure 3 (similar plots for individual diseases are found in Supplementary Material) shows the distribution of the suspected causal agents reported for all SLRD and respective categories communicated to SWORD between 1999 and 2019. The most frequently reported agents across the reporting period were biological substances (42.0%), followed by chemically ill-defined substances (12.6%), and isocyanates (10.1%).

**Figure 3.**
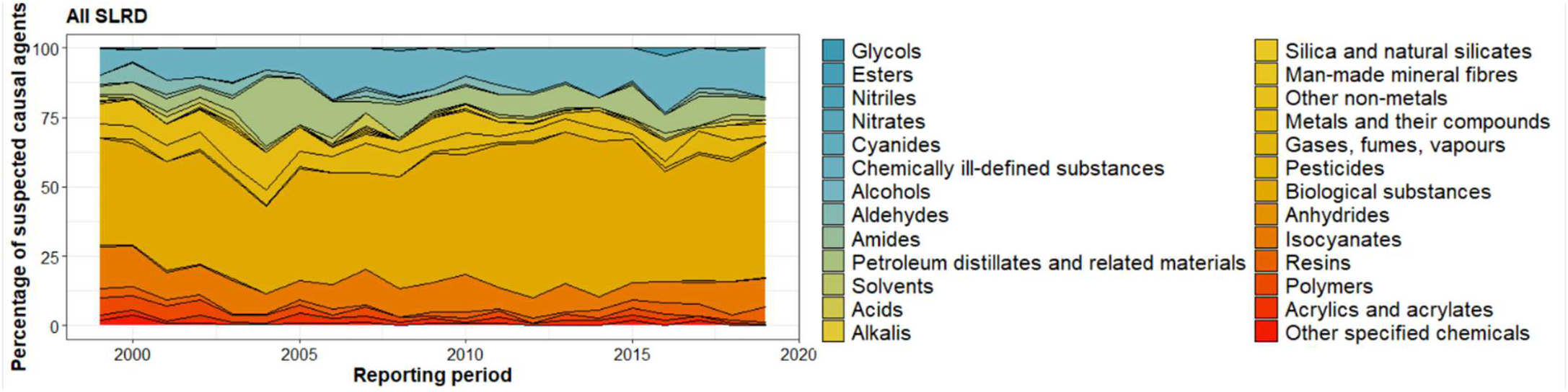
Suspected causal agents of occupational SLRD reported to SWORD (1999-2019). SWORD, Surveillance of Work-Related and Occupational Respiratory Disease.

Finally, other less commonly reported cases included bronchiolitis (9 cases), metal fume fever (9 cases), humidifier fever (8 cases), hyperventilation (8 cases), and cough/laryngitis (4 cases each). More information can be found in Table S1.

## Discussion

This study reports the incidence of SLRD in the UK as communicated by chest physicians to the SWORD surveillance scheme over a recent two-decade period (before the COVID-19 pandemic). Overall case numbers indicated that respiratory diseases with short latency account for approximately 20% of all occupational respiratory disease affecting the UK working population (14) and are comparable to those reported in other European countries (26-29). In previous years in the UK, the annual incidence rate of SLRD has remained stable with moderate decreases in a few disease groups including asthma (30, 14), until the recent reversal of this trend observed after 2014 (31). Reports since the first analyses of SLRD in the UK have kept unchanged regarding the main disease categories (18), however, hypersensitivity pneumonitis cases reported in this study do not seem to be as rare as previously described, and the same is true for cases of rhinitis.

Findings of this study also indicated that the occupations at highest risk of developing SLRD are bakers and flour confectioners and vehicle spray painters (Figure 1). Most SWORD reports for these groups include occupational asthma and rhinitis attributed to flour and diisocyanates (Figure S2 and Figure S3). In the case of asthma, although both substances are responsible for causing sensitisation, they have been shown to differ in a number of ways in terms of properties and disease mechanisms (32). For example, flour is a high molecular weight substance that usually causes IgE-dependent immunologic reactions (33), while diisocyanates are low molecular weight chemicals which are thought to cause asthma by other mechanisms, since specific IgE antibodies to diisocyanates are not observed in the serum of the majority of patients (34). Flour dust has long been recognised as a hazardous substance which can include and combine diverse components used to improve baking, for example, enzymes, amino acids, and chemical additives such as bleaching agents and emulsifiers (35). These substances are potential sensitising agents and are therefore susceptible to cause allergic and non-allergic respiratory reactions. Our data are consistent with the observation that occupational rhinitis in bakers is more common (the highest SRR observed in this study) and usually precedes asthma (9) since approximately 8% of asthma cases reported to SWORD have a co-diagnosis of rhinitis (36, 37).

Exposure to metalworking fluids such as cutting and soluble oils has also been linked to occupational respiratory diseases, due to the inhalation of fluid mists during metal machining processes (38, 39). When considering hypersensitivity pneumonitis, high incidence rates were observed for metal machining setters and setter-operators and metal working machine operatives, even though this lung condition is typically associated with the inhalation of organic dust particles (40, 41). Although mineral oil lubricants are thought to be carcinogenic (42) and cause lipoid pneumonia (43) and pulmonary fibrosis (44), their relationship with SLRD is perhaps less studied. In fact, reports of hypersensitivity pneumonitis due to metalworking fluid aerosols reported before 2000 were relatively uncommon as documented by SWORD and OPRA reports (45) (Figure S4). Several components used in the oil formulation are suspected of causing hypersensitivity pneumonitis including microbial contamination, however, the causal links and disease mechanisms have yet to be investigated (46).

Exposure to microbial organisms in the workplace has been associated with diverse infectious respiratory diseases (17). SWORD reports include infections predominantly caused by bacteria (80%). These are mostly associated with pathogenic mycobacteria and include multidrug-resistant and zoonotic bovine tuberculosis (Figure S1), with most cases observed in healthcare occupations. However, a shift from these pathogens and microorganisms to causal agents such as welding fumes, fungi, moulds and yeast, and ventilation has been observed in recent years (Figure S5). Our results show that nurses have one of the lowest SLRD incidence rates, however, the incidence of infection for these healthcare professionals is significantly raised when compared to all other occupations (Table 2). Nonetheless, according to results of a cohort study based on healthcare workers in the UK, which found that although this occupational group has a higher incidence of tuberculosis than non-healthcare workers, this disease is generally not acquired through occupational exposure (47). Welding trades occupations, on the other hand, are known to be at increased risk of developing pneumococcal pneumonia, in particular lobar pneumonia, which affects one or more lobes of the lung where inflammation and oedema acquire a consolidated pattern. Taking this into consideration, in 2014, the Health and Safety Executive jointly with the manufacturers’ organisation Make UK (formerly known as EEF) and the Cast Metals Federation (CMF) have provided additional guidance on previous advice from the Department of Health issued in 2012, whereby pneumonia vaccination was strongly recommended to employees exposed to welding or metal fumes (48). In this study, we still find welding trades as the occupation with the highest incidence rate of occupational infectious diseases, and it would be interesting to follow up the trend in incidence of pneumonia in welding trades workers after the introduction of this preventive measure.

Clinical diagnosis of occupational SLRD can become challenging when dealing with diseases presenting similar clinical features. SWORD reports of diseases categorised as ‘Other SLRD’ include cases that may have common causes and overlapping symptoms with the otherwise classified subgroups. This is the case for organic dust toxic syndrome and humidifier fever, which include nonimmunologic reactions involving non-specific symptoms.

The SRR used in this study allow for cross-occupational comparisons of incidence rates with all other occupations combined, revealing significant high-risk exposures for SLRD in activities such as bakers and flour confectioners and vehicle spray painters. Although incidence estimates may be improved by adjustments for cases missed due to non-participation of eligible chest physicians and respective response rates (49), we did not take these factors into consideration in this study. However, estimates of variance of total estimated cases due to sample reporting in the numerator were taken into account when calculating confidence intervals. Moreover, potential bias in the LFS-based estimates of the workforce by occupation in the UK is also mitigated by the use of rate ratios.

Considering that SLRD, besides from being preventable, have the ability to significantly improve and even resolve after cessation of work exposure, there are considerable opportunities not just for the prevention of workplace hazards through improved risk assessment but also for better interventions in the populations at risk.

### Key points

- What is already known about this subject Short-latency respiratory disease (SLRD) is very commonly diagnosed worldwide, yet not much is known about the occupations at high risk of developing SLRD in the UK.
- What this study adds This study found significantly raised standardised rate ratios (SRR) for occupational rhinitis, asthma, and hypersensitivity pneumonitis, respectively associated with bakers and flour confectioners, vehicle spray painters, and metal machining operatives.
- What impact this may have on practice or policy Given that SLRD have the potential to improve and even resolve when the occupational hazard is mitigated or removed, the identification of occupational groups and agents associated with SLRD is a promising approach to guide future control and preventive measures.

## Data Availability

All data produced in the present work are contained in the manuscript.

## Acknowledgments

The authors are grateful to all physicians who report to SWORD.

## Supplementary Material

**Table S1.** Diseases specified as ‘Other SLRD’ cases reported to SWORD^*^ as described by the reporters (1999-2019).

| Diagnosis | Number of cases |
| --- | --- |
| Active interstitial pneumonitis | 1 |
| Acute inhalation fever | 1 |
| Adult respiratory | 2 |
| Bronchial pulmonary aspergillosis | 2 |
| Bronchiolitis (including obliterative bronchiolitis) | 9 |
| Cough | 4 |
| Diaphragm palsy | 1 |
| Eosinophilic bronchitis | 2 |
| Hiccups | 1 |
| Humidifier fever | 8 |
| Hyperventilation | 8 |
| Inducible laryngeal obstruction | 2 |
| Invasive Aspergillus sinusitis | 1 |
| Irritant bronchitis | 1 |
| Laryngeal oedema | 1 |
| Laryngitis | 4 |
| Lipoid pneumonia | 1 |
| Metal fume fever | 9 |
| Nasal aspergillosis | 1 |
| Organic dust toxic syndrome | 1 |
| Pleuritis/pneumonitis | 1 |
| Polymer fume fever | 2 |
| Possible alveolitis | 1 |
| Rhinoconjunctivitis | 1 |
| Pulmonary embolus secondary to fracture at work | 1 |
| Sarcoidosis | 2 |
| Sinusitis | 1 |
| Throat irritation and headache | 1 |
| Throat tightness | 1 |
| Toxic bronchitis/pneumonitis | 1 |
| Traumatic pneumothorax | 3 |
| Trichlorethylene hyperventilation | 1 |
| Vocal cord dysfunction | 2 |
| Wheezing | 1 |
| # ribs | 1 |

\*Surveillance of Work-Related and Occupational Respiratory Disease

**Figure S1.**
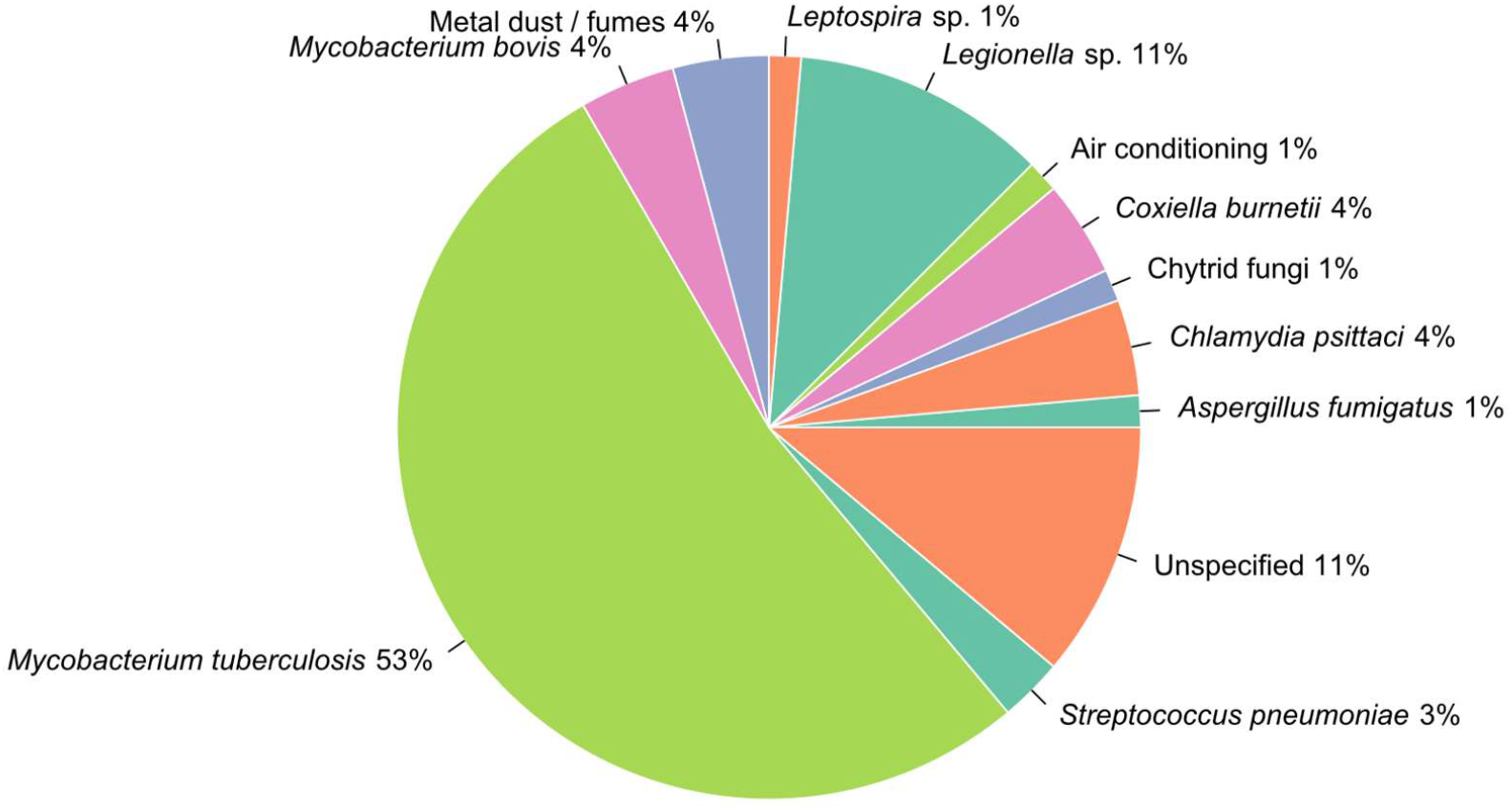
Pathogenic agents of occupational infectious disease reported to SWORD (1999-2019) in the UK. SWORD, Surveillance of Work-Related and Occupational Respiratory Disease.

**Figure S2.**
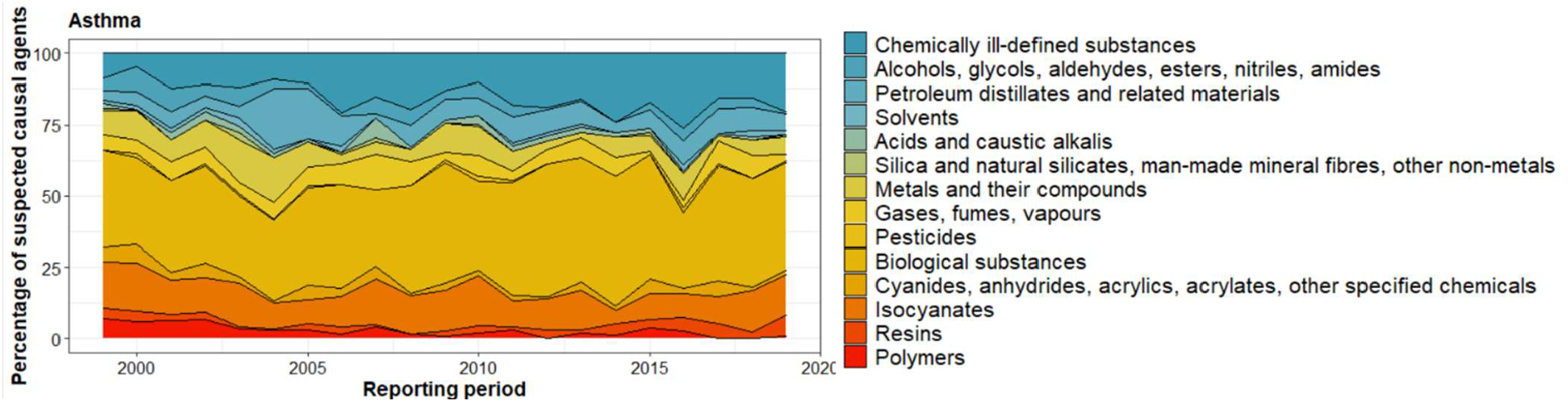
Suspected causal agents of occupational asthma reported to SWORD (1999-2019). SWORD, Surveillance of Work-Related and Occupational Respiratory Disease.

**Figure S3.**
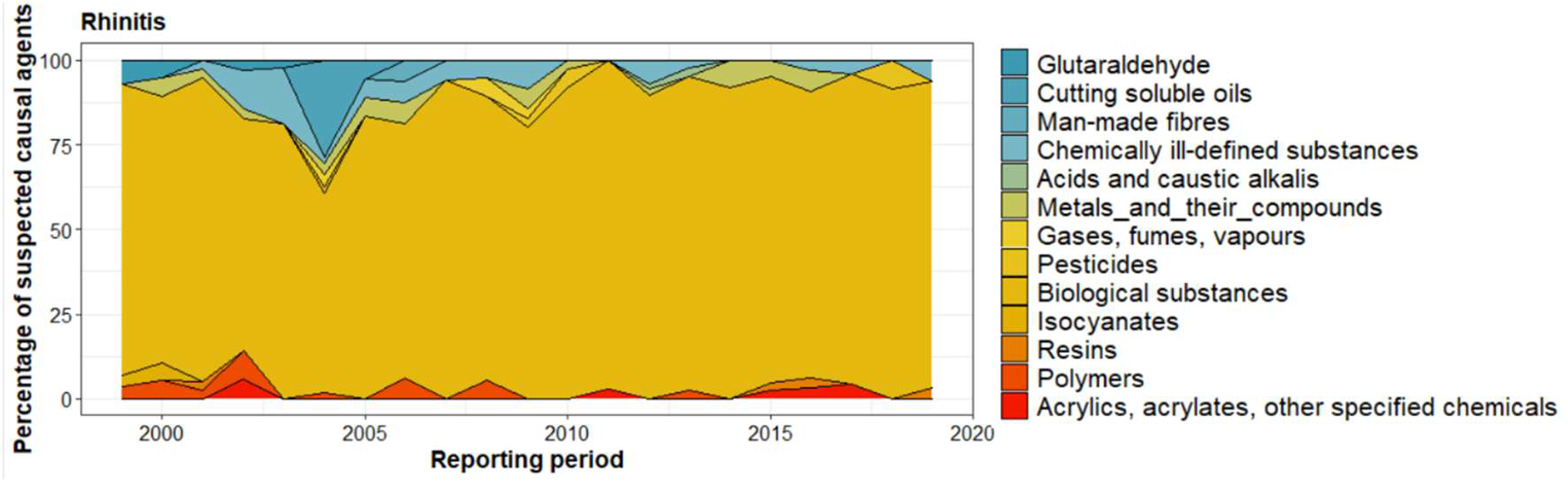
Suspected causal agents of occupational rhinitis reported to SWORD (1999-2019). SWORD, Surveillance of Work-Related and Occupational Respiratory Disease.

**Figure S4.**
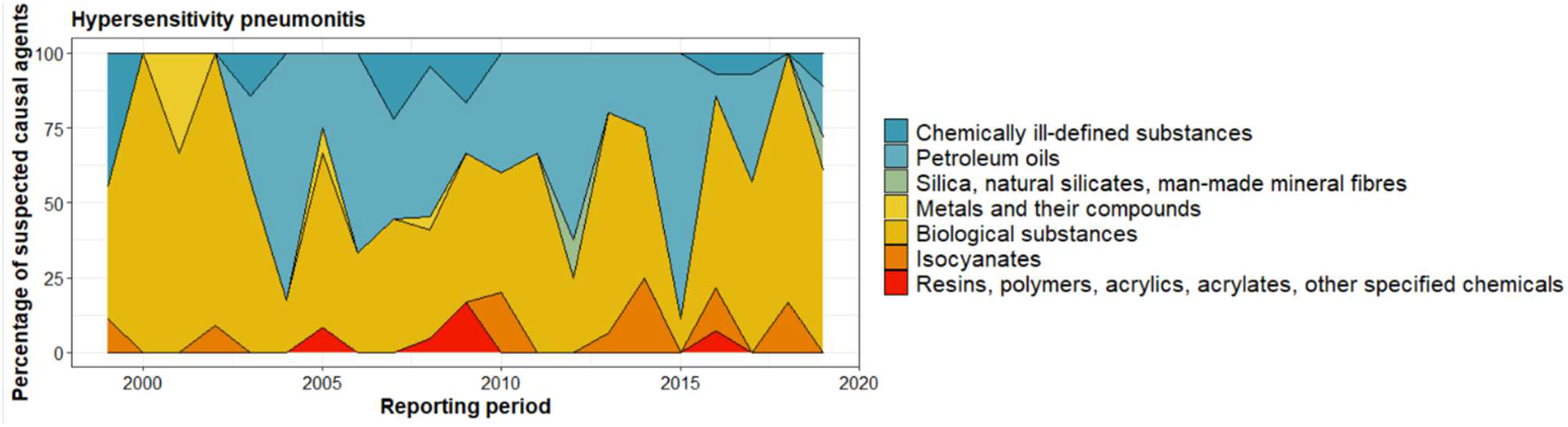
Suspected causal agents of occupational hypersensitivity pneumonitis reported to SWORD (1999-2019). SWORD, Surveillance of Work-Related and Occupational Respiratory Disease.

**Figure S5.**
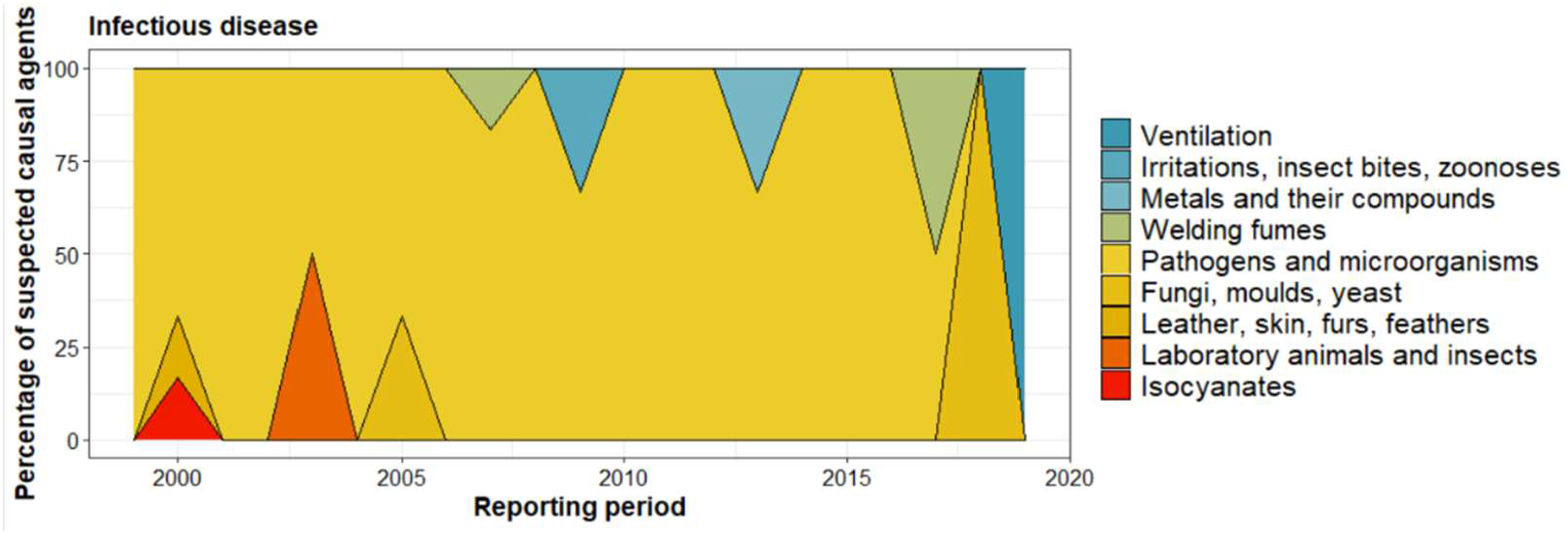
Suspected causal agents of occupational infection reported to SWORD (1999-2019). SWORD, Surveillance of Work-Related and Occupational Respiratory Disease.

